# An Online Program Focusing on Modifiable Lifestyle and Environmental Interventions Improved Pediatric Eczema Symptoms: Results from A Retrospective Observational Study

**DOI:** 10.64898/2026.02.06.26345588

**Authors:** Ana-Maria Temple, Desta Golden, John D. Temple, Christopher R. D’Adamo

**Affiliations:** Integrative Health Carolinas, Charlotte, NC; Global Integrative Health Institute, Charlotte, NC; OvationLab, Richmond, VA; Documenting Hope, Windsor, CT; Department of Family and Community Medicine, University of Maryland School of Medicine, Baltimore, MD

**Keywords:** eczema, pediatric, nutrition, microbiome, stress, environmental toxins

## Abstract

**Background:** Pediatric eczema is a highly prevalent condition that often causes substantial suffering among affected children and their families. Numerous modifiable lifestyle and environmental risk factors for the condition have been identified, although these risk factors and related interventions have generally been studied in isolation. The goal of this study was to evaluate the effects of an integrative program for parents of children with eczema that simultaneously addressed multiple lifestyle and environmental risk factors.

**Methods:** Children with eczema diagnosis who began the online eczema program and provided outcomes data from May 2024 to May 2025 were eligible. The primary outcome was the Patient-Oriented Scoring Atopic Dermatitis (PO-SCORAD), a validated measure of eczema symptoms and burden. Outcomes were assessed at baseline and at one month, two months, and six months after beginning the program. Changes in mean PO-SCORAD scores from baseline throughout the duration of the study were assessed with analysis of variance (ANOVA). Multivariate linear regression modeling of PO-SCORAD scores using population-averaged generalized estimating equations (GEE) were also constructed accounting for baseline PO-SCORAD scores and adjusting for age, sex, presence of any allergy, use of topical corticosteroids, and use of antihistamines.

**Results:** 197 participants were included in the study. The mean baseline PO-SCORAD score was 51.4, which is considered severe eczema. PO-SCORAD scores improved over the course of the study (p<0.0001) and there were statistically significant and clinically meaningful improvements noted after one month (11.3 points, 22.0% improvement), two months (17.8 points, 34.6% improvement), and six months (27.2 points, 52.9% improvement) in the program (p<0.0001). After accounting for baseline PO-SCORAD scores and covariates in regression modeling, there was a 22.5-point (p<0.0001) improvement in PO-SCORAD scores from baseline to final assessment. There was a 31.4-point decrease in PO-SCORAD scores from baseline to final assessment (p<0.0001, 47.2% improvement) among the subgroup of participants with severe eczema symptoms at baseline.

**Conclusions:** An online program focusing on modifiable lifestyle and environmental modifications was associated with clinically meaningful symptom improvements among children with eczema. Symptoms improved relatively quickly and the greatest improvements were noted among children with severe symptoms at baseline.

## Introduction

Eczema has become one of the most common chronic inflammatory skin disorders among children. Current data indicate that approximately 11% of children worldwide suffer from eczema^1^, a prevalence that has risen steadily over the past several decades^2^. Affected children typically present with persistent, relapsing episodes of erythema, intense pruritus, and areas of xerosis. These clinical features typically emerge during infancy and early childhood, adversely impacting sleep quality, school performance, and overall quality of life of both child and family. Epidemiological investigations have revealed a complex interplay between genetic predisposition and environmental triggers. Children with a family history of atopic disease are far more susceptible to developing the disease and lifestyle and environmental influences appear to act as potent accelerants in disease onset and progression^3^,^4^,^5^.

Current medical treatments for pediatric eczema have evolved over the past several decades and can often provide relief. However, there continue to be challenges regarding insufficient resolution of symptoms, adherence, and adverse events in many cases. The standard therapeutic approach typically begins with regular application of emollients to restore and maintain skin barrier integrity, which frequently serves as the foundation upon which other treatments build^6^. For acute inflammatory flares, topical corticosteroids are the first-line medical intervention. These agents are highly effective at reducing inflammation and providing symptomatic relief, with their potency tailored to the severity of the dermatitis and the sensitivity of the affected skin areas. Despite their efficacy, long-term usage of topical corticosteroids carries risks of cutaneous side effects such as skin atrophy, telangiectasia, and striae. Parental concern often leads to underuse or inconsistent application, thereby compromising overall disease control^7^. In cases of moderate to severe eczema that are refractory to topical management, systemic therapies may be deployed, including immunosuppressants and biologic agents. While these therapies have demonstrated promising efficacy, they require careful monitoring due to potential systemic adverse events and variable long-term safety profiles^8^. Collectively, while current pharmacological interventions have significantly improved clinical outcomes for many pediatric patients, treatment failure - whether due to non-adherence, insufficient response, or adverse event-related discontinuation remains a notable challenge that underscores the need for safer and more effective therapeutic strategies.

A growing body of literature has elucidated the myriad non-pharmacological influences that modulate pediatric eczema, including the critical roles of dietary factors, gut microbiota composition, psychological stress, and environmental exposures. Diet plays a pivotal role in skin and immune homeostasis. Early life nutritional factors such as diversified food introduction during infancy^9^, breastfeeding duration^10^, and maternal dietary intake of omega-3 polyunsaturated fatty acids^11^ have all been associated with a reduced risk of eczema development, possibly by modulating both systemic inflammatory responses and the composition of the gut microbiota. Numerous studies have also underscored the importance of the gut-skin axis in pediatric eczema. Dysbiosis in the gut, characterized by reduced microbial diversity and a diminished presence of bacteria that produce short-chain fatty acids, has been linked to impaired immune regulation and increased susceptibility to atopic inflammation^12,13^. Psychological stress can further exacerbate eczema by altering neuroimmune pathways. The release of stress hormones, such as cortisol, may impair skin barrier function and potentiate Th2-skewed inflammatory responses, thereby perpetuating a vicious cycle of flares and pruritus^14,15^. Environmental exposures, including indoor allergens such as house dust mites, pollens, molds, volatile organic compounds from chemical fragrances and cleaning agents, have significant roles in triggering and worsening eczema symptoms^16,17^,18. These exposures not only provoke innate immune responses, but can also disrupt the delicate balance of the skin microbiota, further compromising barrier integrity. Combined, these data suggest that modifiable lifestyle factors are intricately woven into the pathogenesis of pediatric eczema, providing promising targets for interventions that complement and extend beyond traditional pharmacotherapy.

Despite the extensive body of research establishing the roles of modifiable lifestyle factors and environmental exposures in pediatric eczema, there remains a conspicuous paucity of evidence evaluating interactions among these individual risk factors. Furthermore, while systematic reviews and meta-analyses have supported the beneficial effects of targeted probiotic supplementation^19^, dietary modification^20^, allergen avoidance^21^, and stress reduction^22^ in isolation, these investigations have generally focused on single modalities, rather than comprehensively targeting the likely complex interplay among them. The current study aimed to address this critical gap by evaluating outcomes from a comprehensive, multimodality, online program that simultaneously incorporates nutritional guidance, lifestyle modifications, and environmental exposure management for children with eczema.

## Material and Methods

### Study Design

A retrospective observational study of outcomes data from a six-month, online pediatric eczema program emphasizing modifiable lifestyle factors and environmental exposures was conducted. Outcomes data collected from program participants from May 2024-May 2025 were analyzed. Informed consent for analysis and publication of deidentified data was obtained from all program participants. The study was approved by the Institutional Review Board of the Institute for Regenerative and Cellular Medicine (approval # IRCM-2025-448).

### Pediatric Eczema Program

The program under study was an online, multimodal approach to eczema that focuses upon incorporating nutritional interventions, environmental toxin reduction, stress management, and microbiome support to reduce eczema symptom severity.

This six-month program was developed to address the multifactorial nature of eczema by targeting both symptom management and potential root causes. Delivered entirely through a virtual platform, it was accessible to families across geographic boundaries, with structured guidance from a multidisciplinary team of licensed medical doctors, naturopathic doctors, and health coaches. The program included daily support through group sessions, one-on-one consultations, and a moderated community forum to facilitate emotional support.

The program was designed to complement, not replace, standard-of-care dermatologic management for pediatric eczema. Conventional therapies such as topical corticosteroids, calcineurin inhibitors, and emollients were part of the program for reducing acute inflammation and maintaining skin barrier function. By aligning the lifestyle and environmental modification interventions with established medical treatments, the program aimed to improve clinical outcomes and quality of life while reducing reliance on higher-intensity pharmacologic therapy when clinically appropriate.

Areas of program emphasis include:

#### 1. Nutrition and Dietary Modification

Dietary interventions form the cornerstone of the program. Families are guided through the process of reducing processed food intake, limiting added sugars, and replacing common inflammatory triggers. Processed foods, particularly those containing artificial dyes, preservatives, and refined carbohydrates, can exacerbate systemic inflammation and dysbiosis^23,24,25,26^, both of which can often contribute to eczema pathogenesis. Participants are encouraged to replace nutrient-sparse options with whole, minimally processed foods rich in essential fatty acids, antioxidants, vitamins, and minerals. Practical meal plans, shopping guides, and family-friendly recipes are provided to foster long-term adherence and to promote positive dietary habits for the entire household.

#### 2. Nutrient Repletion and Supplementation

Nutrient deficiencies - particularly in vitamin D, zinc, and omega-3 fatty acids - have been implicated in impaired skin barrier function and altered immune responses^27,28,29,30^. The program utilizes individualized nutritional supplementation protocols based on clinical assessment and laboratory evaluation. In addition to correcting suboptimal nutrient levels, targeted supplementation includes probiotics, digestive enzymes, and herbal supplements to support gastrointestinal function and microbial diversity.

#### 3. Gut Health and Microbiome Support

Emerging research highlights the gut-skin axis as a critical factor in eczema development and persistence^31^. This program addresses dysbiosis and impaired gut barrier function by incorporating probiotics with documented benefits for atopic conditions, prebiotic-rich foods, and herbal antimicrobials when appropriate. Digestive support through enzymes and gut-healing nutrients are also utilized to promote microbial balance. Parallel attention is given to the skin microbiome, with the use of specific topical therapies (e.g., hypochlorous acid, vitamin B_12_ cream) to restore microbial diversity and reduce pathogenic colonization, particularly *Staphylococcus aureus*, which is frequently overrepresented in eczema lesions^32^.

#### 4. Stress Management and Emotional Support

Chronic stress can be both a trigger and an amplifier of eczema symptoms, mediated through neuroimmune pathways and behavioral impacts on sleep, scratching, and self-care^14,15,33^. Families in the program receive one-on-one health coaching to address individual stressors and build resilience. Group “healing circles” and live coaching calls create a sense of shared experience and reduce isolation, aimed at empowering both parents and children. Mindfulness practices, breathing exercises, and sleep hygiene strategies are taught and reinforced throughout the program.

#### 5. Environmental Toxin Reduction

Environmental toxins - including household chemicals, dust mites, fragrances, pesticides, and volatile organic compounds - can exacerbate immune dysregulation and contribute to skin barrier breakdown^17,18,34^. Participants are educated on identifying and reducing toxin exposures in their home environments. Practical steps include transitioning to low-toxicity cleaning products, choosing personal care products free of irritants, improving indoor air quality, reducing dust mite exposure and avoiding known contact allergens. These changes are aimed at reducing the overall inflammatory burden on the immune system and support more stable skin health.

The program delivery was designed for interactive engagement. Families had access to daily opportunities for support, including: live group sessions led by clinicians for education and Q&A, one-on-one coaching for individualized care, and a moderated online forum where participants share experiences and receive peer and professional support. This community-centered approach was designed to foster accountability, motivation, and emotional connection, designed to support long-term adherence.

### Study Participants

Children up to 17 years of age with an eczema diagnosis who enrolled in the online program and provided outcomes data from May 2024 to May 2025 were eligible for the study. Participants were included in the analysis if they had completed a baseline outcomes assessment and at least one follow-up outcomes assessment. Exclusion criteria included participants with only one outcomes assessment or those who turned 18 years of age during the year of outcomes data analysis.

### Data Collection

Participant characteristics (e.g., age, sex, allergies, current medical treatments) and outcomes data were collected online via a secure website. The data collected from the platform were stored on an encrypted server. An email or text prompt was sent to each participant’s parent/guardian to provide outcomes data once per month. No incentives were provided to participants for providing their outcomes data.

### Study Outcomes

#### Primary outcome

The primary outcome of the study was the Patient-Oriented Scoring Atopic Dermatitis (PO-SCORAD). The PO-SCORAD is a validated, self-reported measure of eczema symptoms and burden^35,36^. This measure is scored by assessing the following three domains and adding the results to yield a total severity score:

1. Affected surface area: Indicate areas of skin involved by selecting regions of a body diagram. Each area contributes a percentage to the overall skin surface area involved.
2. Intensity of symptoms: Six symptoms (redness, swelling, oozing/crusting, scratch marks, skin thickening, dryness) are each rated on a severity scale from 0 (absent) to 3 (severe).
3. Subjective symptoms: Parents/guardians rate the severity of their child’s itching and sleep loss over the past three days on a scale from 1 (none) to 10 (worst).

These components are summed to produce the total PO-SCORAD score. The composite score allows patients to track eczema severity and changes over time. A change of 8-9 points is considered clinically meaningful^37^.

Secondary outcomes: Secondary outcomes included discontinuation of corticosteroids and other medications and qualitative analysis of parent/guardian feedback.

### Statistical Methods

Descriptive statistics were computed to characterize the study sample. The primary outcomes analysis was a comparison of mean baseline PO-SCORAD score to the final reported score. All program participants who met the inclusion criteria, regardless of time in the eczema program, were included in the primary analysis of the full cohort.

Analysis of variance (ANOVA) was utilized to determine if there were differences in unadjusted mean PO-SCORAD scores across the study period. Tukey’s pairwise t-tests were also utilized to compare unadjusted mean PO-SCORAD scores at baseline to unadjusted mean PO-SCORAD scores at the one-month, two-month, and six-month assessments.

In order to account for potential within-subject correlation in PO-SCORAD scores over time and confounding by participant characteristics, multivariate linear regression modeling of PO-SCORAD scores using population-averaged generalized estimating equations (GEE) was also conducted using PROC GENMOD in SAS. The primary estimate of interest was the difference in the baseline PO-SCORAD assessment to the final PO – SCORAD assessment within each participant. A repeated-measures GEE framework with an exchangeable working correlation structure was used to account for within-subject correlation over time. The baseline to final assessment variable was treated as a categorical fixed effect, with baseline specified as the reference level, such that the model coefficient represented the mean difference in PO-SCORAD scores. Covariates included age, gender, presence of any allergy, use of topical corticosteroids, and use of antihistamines. These variables were selected a priori based on their clinical relevance to eczema severity.

A number of subgroup analyses were also conducted. These included subgroups of participants who provided outcomes data throughout the full six-month program, participants with severe eczema symptoms at baseline (as defined by PO-SCORAD score > 50^38^), participants who discontinued topical steroids, and participants who discontinued antihistamines.

In order to evaluate the potential time in the eczema program required to achieve clinically meaningful improvement, baseline PO-SCORAD scores were also compared to those after one month and two months in the program.

A sensitivity analysis was also conducted to compare baseline characteristics (baseline PO-SCORAD scores, age, sex, allergies, antihistamine use, topical corticosteroid use) of participants who provided outcomes data only at baseline and one month to those who provided data at all four time points under study (baseline, one month, two months, six months).

All statistical analyses were conducted in SAS Version 9.4 (SAS Institute, Cary, NC). Statistical significance was defined as p < 0.05.

## Results

There were 197 participants included in the outcomes analyses of the eczema program. The mean (standard deviation) age of participants was 3.41 (3.40) years and there was a relatively even distribution of participants by sex (51.7% male, 48.3% female). The mean (standard deviation) PO-SCORAD score at baseline was 51.4 (19.0), which falls within the classification of severe eczema.

Allergies were common, with 42.6% of program participants reporting some type of allergy at baseline. Prevalence of specific allergens were as follows: eggs (26.4%), peanuts (24.4%), tree nuts (19.3%), wheat (10.7%), soy (6.1%), shellfish (5.1%), and fish (4.6%). Usage of topical steroids (44.2%) and antihistamines (32.5%) were also common among participants at baseline.

100% (n=197) of participants provided outcomes data at one month, 77.7% (n=153) provided outcomes data at baseline, one month, and two months, and 37.1% (n=73) provided outcomes data at baseline, one month, two months, and six months. Sensitivity analyses revealed that there were no differences (p>0.22) in any baseline participant characteristics (PO – SCORAD scores, age, sex, allergy, antihistamine use, and topical steroid use) between participants who provided outcomes data only at baseline and one month and participants who provided data at all four time points under study (baseline, one month, two months, six months).

The results of the unadjusted analyses comparing PO-SCORAD scores at baseline to the final reported score (full cohort) and after one month, two months, and six months (complete program) are presented in **Table 1**. In brief, ANOVA revealed that there was a statistically significant difference in PO-SCORAD scores across all timepoints in the study (p<0.0001). Tukey’s post-hoc t-tests revealed that there were also statistically significant (p<0.0001) and clinically meaningful (reduction > 8 points) improvements in PO-SCORAD scores from baseline to all subsequent time-points.

**Table 1:** Unadjusted PO-SCORAD Scores over the Course of the Pediatric Eczema Program in the Full Study Sample.

|  | Baseline<br>(n=197) | One<br>month<br>(n=197) | P value | Two<br>months<br>(n=153) | P value | Six<br>months<br>(n=73) | P value | Final<br>score<br>(n=197) | P value |
| --- | --- | --- | --- | --- | --- | --- | --- | --- | --- |
| PO-SCORAD | 51.4<br>(19.0) | 40.1<br>(18.5) | <0.0001 | 33.6<br>(18.1) | <0.0001 | 24.2<br>(14.0) | <0.0001 | 29.4<br>(18.1) | <0.0001 |

The greatest improvements were noted among participants who provided outcomes data at the completion of the six-month program, with a decrease of 31.2 points (56.2% improvement) on the PO-SCORAD. While there was a monotonic improvement in PO-SCORAD scores with greater time participating in the program, there was a clinically meaningful decrease of 11.3 points (22.0% improvement) after just one month.

The results from the covariate-adjusted regression analyses of PO-SCORAD scores are provided in **Table 2**. After adjusting for covariates, the final assessment in the eczema program was associated with a 22.5‐point lower PO‐SCORAD score than baseline assessment (β = -22.5; p < 0.001). Age was positively, but non‐significantly, associated with higher PO-SCORAD scores (β = 0.63 per year; p = 0.061). Sex and allergy status were not associated with PO‐SCORAD scores (p>0.68). Use of topical steroids was associated with lower PO‐SCORAD scores (β = −5.44; p = 0.003) and antihistamine use was associated with higher PO‐SCORAD scores (β = 9.25; p < 0.001).

**Table 2:** Multivariate linear regression model output of PO-SCORAD Scores.

| Predictor<br>(reference category) | $\beta$ (SE) | 95% CI | P |
| --- | --- | --- | --- |
| Final program assessment<br>(first program assessment) | -22.5 (1.43) | -25.3 to -19.7 | <0.001 |
| Age, per year | 0.63 (0.33) | -0.03 to 1.28 | 0.061 |
| Female (male) | -0.90 (2.16) | -5.13 to 3.32 | 0.68 |
| Any allergy: yes (no) | -0.05 (2.03) | -4.03 to 3.93 | 0.98 |
| Topical steroids: yes (no) | -5.44 (1.85) | -9.06 to -1.82 | 0.003 |
| Antihistamines: yes (no) | 9.25 (1.99) | 5.36 to 13.15 | <0.001 |

51.8% (n=102) of participants had severe eczema symptoms at baseline, as reflected by a PO-SCORAD score > 50. There was a 31.4-point decrease in PO-SCORAD scores from baseline assessment to final assessment (p<0.0001, 47.2% improvement) among the subgroup of participants with severe eczema symptoms at baseline. 16% (n=32) of participants discontinued antihistamines during their participation in the program and 11% (n=21) of participants discontinued topical steroids during the program. There was a 15.1-point decrease (32.3% improvement) in PO-SCORAD scores among the subgroup of participants who discontinued antihistamines (p=0.0016) and a 39.3-point decrease (63.4% improvement) in PO-SCORAD scores among the subgroup of participants who discontinued topical steroids (p<0.0001).

## Discussions

This online pediatric eczema program focusing on lifestyle and environmental modifications was associated with statistically significant and clinically meaningful improvements in the PO-SCORAD, a validated measure of eczema symptom severity that is commonly used in clinical practice. While there was a monotonic improvement noted over time in eczema symptom severity throughout the course of the study, clinically meaningful improvements were noted after just one month of the program. The mean baseline PO-SCORAD score of this sample fell within the range considered to be severe symptomatology. Subgroup analysis revealed particularly notable improvements among participants with symptoms that met or exceeded the severe eczema symptoms threshold at baseline, nearly tripling the clinically meaningful improvement threshold.

Collectively, these findings suggest that the improvements in eczema symptoms following lifestyle and environmental modification are generally achieved relatively quickly, particularly among those with severe symptoms. In addition, while the program utilized conventional pharmacotherapy and did not explicitly encourage medication discontinuation, 16% and 11% of participants discontinued their antihistamines and topical steroids, respectively, during the course of the program. Participants who discontinued topical steroids had particularly notable improvements in their PO-SCORAD scores of approximately five-times the clinically meaningful threshold (39-point reduction, 63.4% improvement, p<0.0001). While prospective studies are needed to elucidate the temporal sequence, it is plausible that the considerable improvement that these participants experienced may have obviated the need for topical steroids. While encouraging, longer-term follow-up would be required to better support these hypotheses.

The findings in this study dovetail with a growing body of literature that has revealed that a variety of modifiable lifestyle factors and environmental exposures may contribute to the pathogenesis of pediatric eczema. For instance, reduced microbial exposure in early childhood^39^, and increased consumption of processed foods^40^, exposure to air pollution^34^, mold in the home environment^41^, and volatile organic compounds (VOCs)^42^, phthalates^43^, and bisphenols common in many household cleaning and personal care products^44^ have all individually been implicated in exacerbating skin barrier impairment, dysregulation of the immune system, and association with pediatric eczema.

While the individual contributions have been well-studied and replicated in many populations, the interactions between these risk factors for pediatric eczema are poorly understood. Similarly, while individual lifestyle and environmental modification interventions have shown promise, such as reduction of household VOCs^19^ and nutritional supplementation with vitamin D, probiotics, and prebiotics^45^, there remains a paucity of literature on multimodal interventions aimed at understanding and addressing multiple lifestyle and environmental risk factors. The overarching goal of this study was to begin to address this gap in the literature by evaluating an intervention that simultaneously addressed numerous lifestyle and environmental risk factors for pediatric eczema.

While the improvements in a validated metric of pediatric eczema symptoms noted in this study are encouraging, there are several limitations worthy of mention. First, while all study participants provided outcomes data at baseline and after one month (100%) and most study participants provided outcomes data after two months in the program (77.7%), a relatively smaller percentage (37.1%) of program participants provided data at all time points (baseline, one month, two months, and six months) under study. Loss to follow up was expected given the online nature of outcomes data collection, which is generally more challenging. It is possible that there was also some non-random loss to follow up. However, the sensitivity analysis that was conducted revealed no differences in any participant characteristics at baseline, including baseline eczema symptom severity, which mitigates some of these concerns. The lack of a control arm is another notable limitation of this study. In the absence of a control arm, it is possible that some degree of the improvements noted were due to factors outside of the program. Another limitation of this study is that the feasibility of the combination of lifestyle and environmental interventions themselves outside of the context of a program with coaching and group support cannot be fully established with these data. Further studies would be required to evaluate the effects of a multimodality lifestyle and environmental modification intervention without support. However, the online nature of the coaching and group support featured in the program is relatively resource-efficient and widely scalable, including underserved areas where comprehensive in-person care for pediatric eczema may not be available or feasible.

Further prospective and controlled studies are needed to confirm these findings, but the results of this study suggest that an online program incorporating a multimodal interventional approach combining lifestyle and environmental modifications that have heretofore only been studied in isolation is feasible and can improve the symptoms of pediatric eczema. While the mean improvements on the PO-SCORAD were clinically meaningful after just one month, controlled studies will help establish the effectiveness of lifestyle and environmental modifications relative to other treatment approaches.

## Data Availability Statement

The dataset analyzed in this study has been uploaded to Figshare and is publicly available at 10.6084/m9.figshare.30647390.

## Funding

The research was funded by Integrative Health Courses, Charlotte, NC.

## Conflict of Interest

AMT and JT are owners of Integrative Health Courses. DG and CRD report no conflicts of interest.

## Acknowledgments

The authors would like to acknowledge Amanda Peel for her contributions in the management of the study-related data.

## Notes

### Funding Statement

This study was funded by Integrative Health Courses, Charlotte, NC.

### Author Declarations

The Institutional Review Board of the Institute for Regenerative and Cellular Medicine gave ethical approval for this work.

## References Cited

1. Migliavaca CB, Lazzarini R, Stein C, Escher GN, de Gaspari CN, Dos Santos HWG et al. Prevalence of Atopic Dermatitis: A Systematic Review and Meta-Analysis. Dermatitis. 2024 Aug 12. doi: 10.1089/derm.2024.0165. Epub ahead of print. PMID: 39134072.

2. Langan SM, Mulick AR, Rutter CE, Silverwood R, Asher I, García‐Marcos L, et al. Trends in eczema prevalence in children and adolescents: A Global Asthma Network Phase I Study. Clin Exp Allergy. 2023 Mar;53(3):337–52. doi: 10.1111/cea.14276. Epub 2023 Feb 8. PMCID: PMC10946567.

3. Liu Y, Sun S, Zhang D, Li W, Duan Z, Lu S. Effects of Residential Environment and Lifestyle on Atopic Eczema Among Preschool Children in Shenzhen, China. Front Public Health. 2022 May 16;10:844832. doi: 10.3389/fpubh.2022.844832. PMID: 35651861; PMCID: PMC9149154.

4. Ravn NH, Halling AS, Berkowitz AG, Rinnov MR, Silverberg JI, Egeberg A, Thyssen JP. How does parental history of atopic disease predict the risk of atopic dermatitis in a child? A systematic review and meta-analysis. J Allergy Clin Immunol. 2020 Apr;145(4):1182–1193. doi: 10.1016/j.jaci.2019.12.899. Epub 2019 Dec 28. PMID: 31887393.

5. Standl M, Budu-Aggrey A, Johnston LJ, Elias MS, Arshad SH, Bager P, et al. Gene-Environment Interaction Affects Risk of Atopic Eczema: Population and In Vitro Studies. Allergy. 2025 Aug;80(8):2201–2212. doi: 10.1111/all.16605.

6. Johnson H, Yu J. Current and Emerging Therapies in Pediatric Atopic Dermatitis. Dermatol Ther. 2022 Dec;12(12):2691–2703. doi: 10.1007/s13555-022-00829-4. Epub 2022 Oct 18. PMID: 36258087; PMCID: PMC9674820.

7. Herzum A, Occella C, Gariazzo L, Pastorino C, Viglizzo G. Corticophobia among Parents of Children with Atopic Dermatitis: Assessing Major and Minor Risk Factors for High TOPICOP Scores. J Clin Med. 2023 Oct 27;12(21):6813. doi: 10.3390/jcm12216813. PMID: 37959278; PMCID: PMC10650526.

8. Davari DR, Nieman EL, McShane DB, Morrell DS. Current Perspectives on the Systemic Management of Atopic Dermatitis. J Asthma Allergy. 2021 Jun 1;14:595–607. doi: 10.2147/JAA.S287638. PMID: 34103945; PMCID: PMC8179820.

9. Tuballa A, Connell D, Smith M, Dowsett C, O’Neill H, Albarqouni L. Introduction of allergenic food to infants and allergic and autoimmune conditions: a systematic review and meta-analysis. BMJ Evid Based Med. 2024 Mar 21;29(2):104–113. doi: 10.1136/bmjebm-2023-112445. PMID: 38123975.

10. Kull I, Böhme M, Wahlgren CF, Nordvall L, Pershagen G, Wickman M. Breast-feeding reduces the risk for childhood eczema. J Allergy Clin Immunol. 2005 Sep;116(3):657–61. doi: 10.1016/j.jaci.2005.04.028. PMID: 16159639.

11. Jia Y, Huang Y, Wang H, Jiang H. Effect of Prenatal Omega-3 Polyunsaturated Fatty Acid Supplementation on Childhood Eczema: A Systematic Review and Meta-Analysis. Int Arch Allergy Immunol. 2023;184(1):21–32. doi: 10.1159/000526366. Epub 2022 Oct 14. PMID: 36244339.

12. Forno E, Onderdonk AB, McCracken J, Litonjua AA, Laskey D, Delaney ML, et al. Diversity of the gut microbiota and eczema in early life. Clin Mol Allergy. 2008 Sep 22;6:11. doi: 10.1186/1476-7961-6-11. PMID: 18808715; PMCID: PMC2562383.

13. Pantazi AC, Nori W, Kassim MAK, Balasa AL, Mihai CM, Chisnoiu T, et al. Gut microbiota profile and atopic dermatitis in the first year of life. J Med Life. 2024 Oct;17(10):948–952. doi: 10.25122/jml-2024-0287. PMID: 39720170; PMCID: PMC11665751.

14. Estefan J, Ferreira DC, Cavalcante FS, Dos Santos KRN, Ribeiro M. Investigation of possible relationship between atopic dermatitis and salivary biomarkers, stress, and sleep disorders. World J Clin Cases. 2023 Jun 16;11(17):3958–3966. doi: 10.12998/wjcc.v11.i17.3958. PMID: 37388791; PMCID: PMC10303611.

15. Arndt J, Smith N, Tausk F. Stress and atopic dermatitis. Curr Allergy Asthma Rep. 2008 Jul;8(4):312–7. doi: 10.1007/s11882-008-0050-6. PMID: 18606083.

16. Krupka Olek M, Bożek A, Foks Ciekalska A, Grzanka A, Kawczyk-Krupka A. Assessment of Hypersensitivity to House Dust Mites in Selected Skin Diseases Using the Basophil Activation Test: A Preliminary Study. Medicina (Kaunas). 2024 Oct 1;60(10):1608. doi: 10.3390/medicina60101608. PMID: 39459395; PMCID: PMC11509700.

17. Kim K. Influences of Environmental Chemicals on Atopic Dermatitis. Toxicol Res. 2015 Jun;31(2):89–96. doi: 10.5487/TR.2015.31.2.089. PMID: 26191377; PMCID: PMC4505354.

18. Luschkova D, Zeiser K, Ludwig A, Traidl-Hoffmann C. Atopic eczema is an environmental disease. Allergol Select. 2021 Aug 23;5:244–250. doi: 10.5414/ALX02258E. PMID: 34476334; PMCID: PMC8383845.

19. Kim K, Lee E, Kim M, Lee KS, Sol IS, Min TK, et al. Therapeutic effectiveness of probiotics for atopic dermatitis: A systematic review and meta-analysis of randomized controlled trials with subgroup analysis. Asian Pac J Allergy Immunol. 2025 Sep;43(3):428–438. doi: 10.12932/AP-280323-1576. PMID: 37578483.

20. Oykhman P, Dookie J, Al-Rammahy H, de Benedetto A, Asiniwasis RN, LeBovidge J, et al. Dietary Elimination for the Treatment of Atopic Dermatitis: A Systematic Review and Meta-Analysis. J Allergy Clin Immunol Pract. 2022 Oct;10(10):2657-2666.e8. doi: 10.1016/j.jaip.2022.06.044. Epub 2022 Jul 19. PMID: 35987995.

21. Yepes-Nuñez JJ, Guyatt GH, Gómez-Escobar LG, Pérez-Herrera LC, Chu AWL, Ceccaci R, et al. Allergen immunotherapy for atopic dermatitis: Systematic review and meta-analysis of benefits and harms. J Allergy Clin Immunol. 2023 Jan;151(1):147–158. doi: 10.1016/j.jaci.2022.09.020. Epub 2022 Sep 30. PMID: 36191689.

22. Yosipovitch G, Canchy L, Ferreira BR, Aguirre CC, Tempark T, Takaoka R, et al. Integrative Treatment Approaches with Mind-Body Therapies in the Management of Atopic Dermatitis. J Clin Med. 2024 Sep 11;13(18):5368. doi: 10.3390/jcm13185368. PMID: 39336855; PMCID: PMC11432615.

23. Chassaing, B, Compher, C, Bonhomme, B, Liu, Q, Tian, Y, Walters, et al. Randomized Controlled-Feeding Study of Dietary Emulsifier Carboxymethylcellulose Reveals Detrimental Impacts on the Gut Microbiota and Metabolome. Gastroenterology 2022, 162, 743–756.

24. Chassaing, B, van de Wiele, T, de Bodt, J, Marzorati, M, Gewirtz, A.T. Dietary emulsifiers directly alter human microbiota composition and gene expression ex vivo potentiating intestinal inflammation. Gut 2017, 66, 1414–1427.

25. Cuevas-Sierra, A, Milagro, F.I, Aranaz, P, Martínez, J.A, Riezu-Boj, J.I. Gut Microbiota Differences According to Ultra-Processed Food Consumption in a Spanish Population. Nutrients 2021, 13, 2710.

26. Rondinella D, Raoul PC, Valeriani E, Venturini I, Cintoni M, Severino A et al. The Detrimental Impact of Ultra-Processed Foods on the Human Gut Microbiome and Gut Barrier. Nutrients. 2025 Feb 28;17(5):859. doi: 10.3390/nu17050859. PMID: 40077728; PMCID: PMC11901572.

27. Ahmed Mohamed A, Salah Ahmed EM, Farag YMK, Bedair NI, Nassar NA, Ghanem AIM. Dose-response association between vitamin D deficiency and atopic dermatitis in children, and effect modification by gender: a case-control study. J Dermatolog Treat. 2021 Mar;32(2):174–179. doi: 10.1080/09546634.2019.1643447. Epub 2019 Aug 2. PMID: 31296076.

28. Wang SS, Hon KL, Kong AP, Pong HN, Wong GW, Leung TF. Vitamin D deficiency is associated with diagnosis and severity of childhood atopic dermatitis. Pediatr Allergy Immunol. 2014 Feb;25(1):30–5. doi: 10.1111/pai.12167. Epub 2014 Jan 3. PMID: 24383670.

29. Gray NA, Dhana A, Stein DJ, Khumalo NP. Zinc and atopic dermatitis: a systematic review and meta-analysis. J Eur Acad Dermatol Venereol. 2019 Jun;33(6):1042–1050. doi: 10.1111/jdv.15524. Epub 2019 Mar 15. PMID: 30801794.

30. Niseteo T, Hojsak I, Ožanić Bulić S, Pustišek N. Effect of Omega-3 Polyunsaturated Fatty Acid Supplementation on Clinical Outcome of Atopic Dermatitis in Children. Nutrients. 2024 Aug 24;16(17):2829. doi: 10.3390/nu16172829. PMID: 39275147; PMCID: PMC11397185.

31. Yang L, Xia JN. Beyond the Skin: Exploring the Gut-Skin Axis and Metabolic Pathways in Atopic Dermatitis Pathogenesis. Int J Gen Med. 2025 Oct 8;18:6123–6136. doi: 10.2147/IJGM.S550152. PMID: 41084738; PMCID: PMC12515402.

32. Ogonowska P, Gilaberte Y, Barańska-Rybak W, Nakonieczna J. Colonization with Staphylococcus aureus in Atopic Dermatitis Patients: Attempts to Reveal the Unknown. Front Microbiol. 2021 Jan 11;11:567090. doi: 10.3389/fmicb.2020.567090. PMID: 33505363; PMCID: PMC7830525.

33. Lin TK, Zhong L, Santiago JL. Association between Stress and the HPA Axis in the Atopic Dermatitis. Int J Mol Sci. 2017 Oct 12;18(10):2131. doi: 10.3390/ijms18102131. PMID: 29023418; PMCID: PMC5666813.

34. Dijkhoff IM, Drasler B, Karakocak BB, Petri-Fink A, Valacchi G, Eeman M et al. Impact of airborne particulate matter on skin: a systematic review from epidemiology to in vitro studies. Part Fibre Toxicol. 2020 Jul 25;17(1):35. doi: 10.1186/s12989-020-00366-y. PMID: 32711561; PMCID: PMC7382801.

35. Severity scoring of atopic dermatitis: the SCORAD index. Consensus Report of the European Task Force on Atopic Dermatitis. Dermatology. 1993;186(1):23–31. doi: 10.1159/000247298. PMID: 8435513.

36. Stalder JF, Barbarot S, Wollenberg A, Holm EA, De Raeve L, Seidenari S, et al. PO-SCORAD Investigators Group. Patient-Oriented SCORAD (PO-SCORAD): a new self-assessment scale in atopic dermatitis validated in Europe. Allergy. 2011 Aug;66(8):1114–21. doi: 10.1111/j.1398-9995.2011.02577.x. Epub 2011 Mar 18. PMID: 21414011.

37. Schram ME, Spuls PI, Leeflang MM, Lindeboom R, Bos JD, Schmitt J. EASI, (objective) SCORAD and POEM for atopic eczema: responsiveness and minimal clinically important difference. Allergy. 2012 Jan;67(1):99–106. doi: 10.1111/j.1398-9995.2011.02719.x. Epub 2011 Sep 27. PMID: 21951293.

38. Kisieliene I, Aukstuolyte B, Mainelis A, Rudzevicienė O, Bylaite-Bucinskiene M, Wolenberg A. Associations between Atopic Dermatitis and Behavior Difficulties in Children. Medicina (Kaunas). 2024 Mar 17;60(3):492. doi: 10.3390/medicina60030492. PMID: 38541218; PMCID: PMC10972345.

39. Chatenoud L, Bertuccio P, Turati F, Galeone C, Naldi L, Chatenoud L, et al; HYGIENE Study Group. Markers of microbial exposure lower the incidence of atopic dermatitis. Allergy. 2020 Jan;75(1):104–115. doi: 10.1111/all.13990. Epub 2019 Aug 12. PMID: 31321780.

40. Berni Canani R, Carucci L, Coppola S, D’Auria E, O’Mahony L, Roth-Walter F et al. Ultra-processed foods, allergy outcomes and underlying mechanisms in children: An EAACI task force report. Pediatr Allergy Immunol. 2024 Sep;35(9):e14231. doi: 10.1111/pai.14231. PMID: 39254357.

41. Zhang R, Weschler LB, Ye J, Wang Z, Deng Q, Li B, et al. Associations between home environmental factors and childhood eczema and related symptoms in different cities in China. Heliyon. 2023 Nov 8;9(11):e21718. doi: 10.1016/j.heliyon.2023.e21718. PMID: 38027650; PMCID: PMC10661510.

42. Kwon JH, Kim E, Chang MH, Park EA, Hong YC, Ha M, et al. Indoor total volatile organic compounds exposure at 6 months followed by atopic dermatitis at 3 years in children. Pediatr Allergy Immunol. 2015 Jun;26(4):352–8. doi: 10.1111/pai.12393. PMID: 25868723.

43. Zhang H, Chen S, Chen X, Zhang Y, Han Y, Li J, et al. Exposure to phthalate increases the risk of eczema in children: Findings from a systematic review and meta-analysis. Chemosphere. 2023 Apr;321:138139. doi: 10.1016/j.chemosphere.2023.138139. Epub 2023 Feb 13. PMID: 36791818.

44. Kim EH, Jeon BH, Kim J, Kim YM, Han Y, Ahn K, et al. Exposure to phthalates and bisphenol A are associated with atopic dermatitis symptoms in children: a time-series analysis. Environ Health. 2017 Mar 9;16(1):24. doi: 10.1186/s12940-017-0225-5. PMID: 28274229; PMCID: PMC5343323.

45. Vassilopoulou E, Comotti A, Douladiris N, Konstantinou GN, Zuberbier T, Alberti I, et al. A systematic review and meta-analysis of nutritional and dietary interventions in randomized controlled trials for skin symptoms in children with atopic dermatitis and without food allergy: An EAACI task force report. Allergy. 2024 Jul;79(7):1708–1724. doi: 10.1111/all.16160. Epub 2024 May 23. PMID: 38783644.

